# Users’ perception of the OH-EpiCap evaluation tool based on its application to nine national antimicrobial resistance surveillance systems

**DOI:** 10.1101/2023.03.15.23287323

**Authors:** Pedro Moura, Lucie Collineau, Marianne Sandberg, Laura Tomassone, Daniele De Meneghi, Madelaine Norström, Houda Bennani, Barbara Häsler, Mélanie Colomb-Cotinat, Clémence Bourély, Maria-Eleni Filippitzi, Sarah Mediouni, Elena Boriani, Muhammad Asaduzzaman, Manuela Caniça, Cécile Aenishaenslin, Lis Alban

## Abstract

Antimicrobial resistance (AMR) surveillance systems involve multiple stakeholders and multilevel standard operating procedures, which increase in complexity with further integration of the One Health (OH) concept. AMR is a OH challenge. It is crucial for the success of an AMR surveillance system to evaluate its performance in meeting the proposed objectives, while complying with resource restrictions. The OH-EpiCap tool was created to evaluate the degree of compliance of hazard surveillance activities with essential OH concepts across there dimensions: organization, operational activities, and impact of the OH surveillance system.

To present feedback on the application of the OH-EpiCap from a user’s perspective, the tool was used to evaluate nine national AMR surveillance systems, each with different monitoring contexts and objectives. The OH-EpiCap tool was assessed using the updated CoEvalAMR methodology. This methodology evaluates the content themes and functional aspects of the tool in a standardized way, while it also captures the user’s subjective experiences in using the tool via a strengths, weaknesses, opportunities, and threats (SWOT) approach.

The results of the evaluation of the OH-EpiCap are presented and discussed. The OH-EpiCap is an easy-to-use tool, which can facilitate a fast macro-overview of the application of the OH concept to a surveillance activity, when used by specialists in the matter, serving as a basis for the discussion of possible adaptations of AMR surveillance activities, or targeting areas that may be further investigated using other pre-established tools.

## 1 Introduction

The World Health Organization (WHO), the Food and Agriculture Organization of the United Nations (FAO), the World Organisation for Animal Health (WOAH) and The United Nations Environment Programme (UNEP) have established the One Health (OH) High Level Expert Panel (OHHLEP) (1). This Panel defines OH as “an integrated, unifying approach that aims to sustainably balance and optimize the health of people, animals, and ecosystems” recognizing that the health of humans, domestic and wild animals, plants, and the wider environment are closely linked and interdependent (1).

The circulation of microorganisms carrying AMR genes cannot be restricted to one specie or sector, or to a specific geographical location. Also, given the cross use of certain antimicrobial agents in humans, animals, and plants, AMR is one of the quintessential examples of a global scale OH challenge (2). Therefore, a coordinated, multisectoral and multidisciplinary approach is necessary to address the issue (3,4).

Integrated surveillance, according to Aenishaenslin et al., is the “systematic collection, analysis, interpretation of data, and dissemination of information collected from different components of a system to provide a global, multidisciplinary, multi-perspective understanding of a health problem and to inform system-based decisions”(5). These actions should be coordinated between the human, animal and environmental sectors (6).

The application of this concept to national surveillance systems is essential to better understand AMR genes emergence and dispersion and to sustain risk mitigation decisions (7). A strategic framework, which supports intersectoral collaboration in national strategies against AMR, aiming to keep antimicrobials (AM) effective for future generations of people and animals, has recently been released by the OHHLEP (8).

Technological innovations and new health challenges may arise and impact a system’s performance and demands. Therefore, conducting regular evaluations of a surveillance system’s processes and performance is crucial to assess if the established objectives are being met in the most cost-efficient way (9). OH initiatives should preferably be evaluated using a methodology that targets all disciplines encompassed and estimate the potential added value of the current approach over a less integrated one (10). The objectives of the evaluation should be made clear from the start, and an overview of the systems’ surveillance components should be produced to guide it, and to balance the objectives of the evaluation with the available resources to perform it (9).

The international network CoEvalAMR was established in 2019 with the aim of providing guidance to help users in choosing an assessment tool from a catalogue of tools available to evaluate antimicrobial use (AMU) and AMR surveillance systems (11). Moreover, it was the aim to guide future applications and improvement of the tools assessed and the development of new tools. To meet all these aims, a methodology focusing on the users’ perception of the tool was developed in Phase 1 of the CoEvalAMR network (12). The methodology has recently been updated and further refined as part of the work undertaken in Phase 2 of the network. It encompasses the evaluation of descriptive and functional aspects, together with an assessment of content themes and SWOT questions (13). The original methodology has previously been used by Sandberg et al. to provide feedback on six different evaluation tools based on their application in eight countries (12).

The OH-EpiCap tool has been developed by the MATRIX consortium, funded by the One Health European Joint Program (14) to systematize the characterization of epidemiological surveillance activities in a national surveillance system. More specifically, the purpose of the OH-EpiCap is to facilitate the evaluation and reinforcement of national capacities and capabilities for OH integrated surveillance of zoonotic hazards (14). In the present work, users’ feedback was provided on the OH-EpiCap, which was selected because it is a new tool that is presented as an easy-to-apply tool, covering previously overlooked aspects such as the impact of integrated surveillance.

The objectives of this work were to:

i. Apply and evaluate the OH-EpiCap tool using the updated CoEvalAMR user’s perception methodology
ii. Present feedback on the application of the OH-EpiCap tool to nine national AMR surveillance systems, with different monitoring contexts and objectives.

## 2. Materials and methods

### 2.1 The OH-EpiCap tool

The OH-EpiCap tool is composed of three thematic domains (called dimensions), each with four different targets that are again segmented into four indicators, leading to a total of 48 standardized indicators, briefly presented in Table 1. These indicators are presented in the form of a questionnaire containing single choice questions with five options to choose from, to be answered using a semi-quantitative scale from 1 to 4, with 4 representing the best scenario for integrated OH surveillance. If a certain indicator is not applicable to the system under evaluation, it is possible to select “non-applicable” as an answer. All indicators in each target area have the same weight and the average value of the answers given is converted into the score of the target area. The tool also includes a graphical interface where the results of the evaluation are presented in a dashboard that can be exported as a report. The OH-EpiCap tool is available at the website: https://freddietafreeth.shinyapps.io/OH-EpiCap/.

**Table 1:** Dimensions, targets and indicators evaluated by the OH-EpiCap tool – modified after (15)

| **<u>Dimension 1: Organisation</u>** |  |  |  |
| --- | --- | --- | --- |
| **Target 1.1 Formalisation:** common aim, support documentations, shared leadership, and definition of roles/composition of coordination committees | **Target 1.2 Coverage:** inclusion of all relevant actors, disciplines, sectors, geography, populations, and related hazards | **Target 1.3 Resources:** budget and human resources, program training, and sharing of resources | **Target 1.4 Evaluation and resilience:** internal and external evaluations, development/ implementation of corrective measures, and adaptability to change |
| **<u>Dimension 2: Operational activities</u>** |  |  |  |
| **Target 2.1 Data collection and methods sharing:** multisectoral collaboration in the design of surveillance protocols and data collection, harmonization of laboratory techniques and data warehousing | **Target 2.2 Data sharing:** data sharing agreements, assessment of data quality, usefulness of shared data, and the compliance of data with the FAIR (findability, accessibility, Interoperability and Reusability) principle | **Target 2.3 Data analysis and interpretation:** multisectoral integration for data analysis, sharing of analysis techniques, sharing of scientific expertise, and harmonization of indicators | **Target 2.4 Communication:** internal and external communication, dissemination to decision-makers, and information sharing in case of suspicion/ particular events |
| **<u>Dimension 3: Impact</u>** |  |  |  |
| **Target 3.1 Technical outputs:** timely detection of emergence, epidemiological knowledge improvement, increased effectiveness of surveillance, and reduction of operational costs | **Target 3.2 Collaborative added value:** strengthening of the OH team and network, international collaboration, and common strategy (road map) design | **Target 3.3 Immediate and intermediate outcomes:** advocacy, awareness, preparedness, and interventions based on the information generated | **Target 3.4 Ultimate outcomes:** research opportunities, policy changes and behavioural changes and better health outcomes |

### 2.2 Data collection

The number of individuals involved in the evaluation of each case study varied from one to five; these individuals are referred to as “assessors” throughout the text. The assessors filled in the OH-EpiCap evaluation tool during either a one or two round workshop that lasted a total of two to eight hours. All the assessors involved had expertise in AMR surveillance in the country they represented for this study, answering the indicator questions according to their own work experience or knowledge from previous activities. Whenever needed, additional experts and information sources were consulted. The number of assessors and their affiliation, the type of workshop conducted, and the total duration of the evaluation are described for each country case study in Table S1. The surveillance system including its main aims evaluated in each country case study can be consulted in Table 2. The systems were selected by the assessors for their own convenience and interest.

**Table 2:** National AMR surveillance systems evaluated using the OH-EpiCap tool.

| Country | Name of the system | Main aims of the system |
| --- | --- | --- |
| Bangladesh | One Health Event Based Surveillance (EBS) | • Develop a ‘One Health surveillance system platform’ to enable early detection of disease outbreaks. Coordinated joint response to disease outbreaks |
| Belgium | AMR-AMU surveillance program in the context of developing the OH AMU-AMR national report (OH belmap) | • Summarize results and trends of existing monitoring programs: related to the consumption of antibiotic agents for food animals and humans and to the monitoring occurrence of antimicrobial resistance in bacteria isolated from food animals, humans and food of animal origin<br>• Identify blind spots in monitoring programs and make recommendations to improve future monitoring |
| Canada | Canadian Integrated Program for Antimicrobial Resistance Surveillance (CIPARS) | • Provide an integrated approach to monitor trends in antimicrobial resistance and antimicrobial use in humans & animals and help identify appropriate measures to contain the emergence and spread of resistant bacteria between animals, food, and people in Canada<br>• Facilitate assessment of the public health impact of antimicrobials used in humans & agriculture to support the creation of evidence-based policies to control AMU in hospital, community, and agricultural settings<br>• Provide timely analysis and dissemination of surveillance data to stakeholders, and facilitate knowledge translation via targeted communications products<br>• Allow accurate comparisons with other countries that use similar surveillance systems (NARMS, DANMAP)<br>• Provision of data for Health Canada—Veterinary Drugs Directorate for new antimicrobial drug approval processes and post-approval monitoring |
| Denmark | Danish Program for surveillance of antimicrobial consumption and resistance in bacteria from food animals, food and humans (DANMAP) | • Monitor the consumption of antimicrobial agents for food animals and humans and the occurrence of antimicrobial resistance in bacteria isolated from food animals, food of animal origin and humans<br>• Study associations between antimicrobial consumption and antimicrobial resistance<br>• Identify routes of transmission and areas for further research studies |
| France | Surveillance system for AMR, AMU and antimicrobial residues | • Monitor trends of AMU and AMR in humans and animals, incl. in diseased animals<br>• Assess what is common to several sectors and what is not<br>• Inform policy recommendations and assess the impact of interventions |
| Italy | ClassyFarm | • Risk categorization of farms according to an integrated approach containing biosecurity, welfare, AMU/AMR, animal health and lesions at slaughterhouse |
| Norway | The surveillance programme for antimicrobial resistance in human pathogens (NORM) and the monitoring programme for antimicrobial resistance in bacteria from feed, food and animals (NORM-VET) | NORM:<br>• Collect and process data about antibiotic resistance of microbe isolates to determine the incidence and prevalence of antibiotic resistance and monitor changes over time<br>• Drive, promote and provide a basis for research to understand why microbes develop antibiotic resistance, with a view to promoting and developing preventive measures in the treatment of infectious diseases<br>• Provide a basis to give health advice and information on measures that could prevent development antimicrobial drug resistance to the public and local, regional and central health authorities<br>• Give the Norwegian health authorities a foundation to contribute to international statistics within specific areas<br>NORM-VET:<br>• Provide and present data on the occurrence and distribution of antimicrobial resistance over time.<br>• Describe the relationship between the use of antimicrobials and occurrence of resistance in the veterinary and food production sectors.<br>• The information generated is used for research, setting policies, assessing risks, and evaluating interventions |
| Portugal | Infection Prevention and Control and Antimicrobial Resistance Program (PPCIRA) | • Monitor the occurrence of antimicrobial resistance in bacteria isolated from humans<br>• Identify routes of transmission<br>• Detect and monitor outbreaks caused by bacteria with antimicrobial resistant genes<br>• Prevent the emergence and transmission of bacteria with antimicrobial resistant genes |
| United Kingdom | Surveillance system for AMU and AMR in the UK | • Monitor AMU in humans and animals<br>• Monitor trends of AMR in bacteria isolated from humans, food producing animals, and food of animal origin<br>• Detect new and emerging AMR threats<br>• Inform policy recommendations and assess the impact of interventions |

### 2.3 Data analysis

The users’ perception methodology used to evaluate the OH-EpiCap was developed during Phase 1 of the CoEvalAMR network project and was described in Sandberg et al., (12). The original methodology consisted of questions related to 1) functional aspects, 2) content themes as well as 3) strengths, weaknesses, opportunities and threats (SWOT). The methodology has recently been updated and further refined based among others on experience using the methodology (13). As stated above, it was created in the scope of the CoEvalAMR network to systematically capture the user’s experience when assessing an integrated system for AMR surveillance using a pre-designed evaluation tool. The updated version consists of the following components: 1) A general description of the case study and the tool, 2) two standardized scoring schemes, one regarding functional aspects, another for content themes, and 3) a SWOT analysis, as described below. The OH-EpiCap tool was evaluated by the assessors using the methodology, and the outcomes of evaluation were summarized and presented in the Results section.

The functional aspects of the OH-EpiCap were scored semi-quantitatively using a scale from 1-4 or “non-applicable”. Groups composed of several functional aspects were averaged in each evaluation case. The median, maximum and minimum of the scores given by the assessors are presented in a radar diagram in Figure 1A of the results section. The content themes of the OH-EpiCap were also scored semi-quantitatively using a scale from 1-4. The three segments that compose the theme “Integration” were averaged in each evaluation case, and the median, maximum and minimum of the scores given by the assessors are presented in a radar diagram in Figure 1B of the results section.

**Figure 1:**
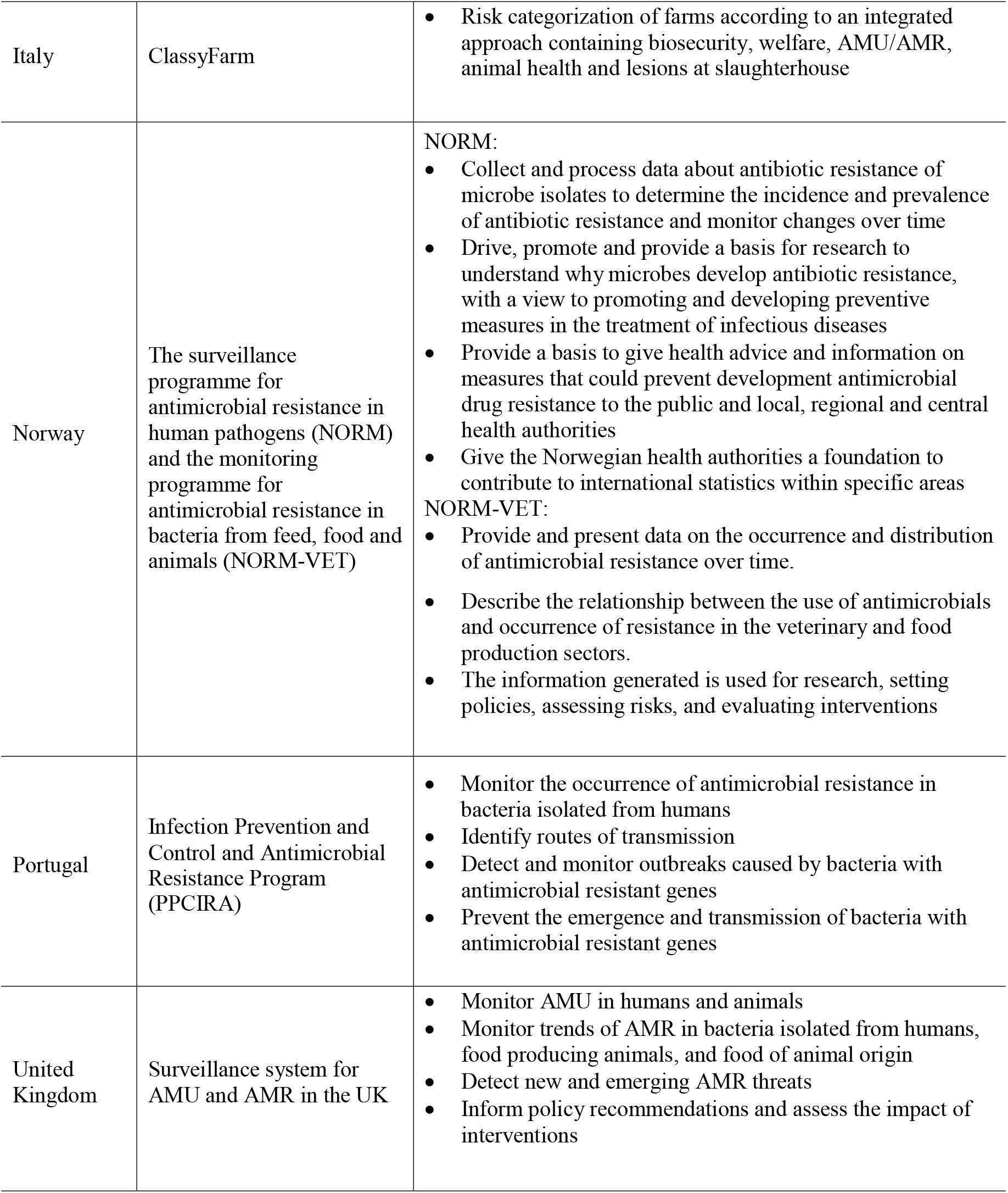
Evaluation of the functional aspects (1A) and content themes (1B) of the OH-EpiCap tool according to the CoEvalAMR user’s perception methodology based upon nine case studies.

The SWOT analysis was undertaken to capture the assessors’ subjective experiences when applying the OH-EpiCap. Hence, assessors were asked to report the strengths, weaknesses, opportunities and threats (SWOT) of the evaluation tool. More specifically, the following wording accompanied each component: Strengths: “The strengths of this tool are”, Weaknesses: “The weaknesses of this tool are”, Opportunities: “The added value(s) of using this tool is” and Threats: “This tool might be criticized because of”. A qualitative synthesis of the feedback provided by the assessors was performed following the same principles as described by Sandberg et al. (2021), which were based on grounded theory (16): all individual sentences were collected, after this, sentences with similar content were simplified and condensed into one sentence. The synthesis was performed by three of the authors and later verified by the assessors and it can be found in Table 4.

## 3 Results

The products of the evaluation conducted by the assessors using the updated CoEvalAMR user’s perception methodology are presented below, segmented into the following: Functional aspects (Figure 1A), Content themes (Figure 1B) and SWOT analysis (Table 3)

**Table 3.** Outcome of SWOT analysis of the OH-EpiCap tool, based on an application of the tool in nine country cases.

| Topic and meaning | Synthesis of the comments provided |
| --- | --- |
| <u>Strengths</u><br>The strengths of this tool are | A feasible compromise between comprehensiveness in quantity of information captured and human/time resources required to carry out evaluation.<br>Simple and well-organized design, following a user-friendly step-by-step approach with boxes to check.<br>No previous extensive training is needed to use it.<br>The provided glossary encompassing explanations of what is meant by an expression is very helpful and increases the ease and swiftness of use.<br>Produces visually attractive figures, encompassed in a report, which provide a good overview of the answers given and make it easy to share and communicate the results. An example of which can be seen in Figure S1 of the Supplementary material.<br>In the report, general suggestions for further improvements and indicators of good adherence to OH principles are provided.<br>It is available for free, useful for single or multidisciplinary settings and suitable for any country.<br>If the people who have a deep understanding of the surveillance system and all the main processes are consulted, it produces a lot of food for thought. |
| <u>Weaknesses</u><br>The weaknesses of this tool are | Some of the indicators could be further simplified, to facilitate their interpretation.<br>Although comprehensive, the evaluation products are superficial, and they cannot be directly translated into action, requiring further investigation;<br>If surveillance initiatives are based on one dominating OH pillar, it is not easy to answer some indicators which are structured to catch multi-sectoral/disciplinary collaborations.<br>Some criteria are difficult to score without dedicated ad-hoc studies.<br>Sometimes difficult to delineate which impacts comes from OH surveillance vs sectoral surveillance (Dimension 3).<br>Some indicators aiming at evaluating effectiveness refer more to technical performance of surveillance (sensitivity, timeliness) vs its capacity to inform decision-making.<br>The tool is sometimes hard to apply to a system which integrates data from multiple domains such as AMR and AMU in animals and humans, but is managed by only one institution, as several items refer to inter-institutions collaboration and governance. |
| <u>Opportunities</u><br>The added value(s) of using this tool is | Helpful to identify new areas that should be further investigated and to initiate discussion around the possibility of adapting the surveillance systems.<br>Provides a good overview of a surveillance system targeting one hazard, or a component of a complex system.<br>Evaluation can be performed in a short time, so it may be done frequently, and after relevant updates.<br>Provides an evaluation at a macroscopic scale of the overall “OH-ness” of the system and facilitates an overall description of the system.<br>Can be used pragmatically for preliminary assessment.<br>Useful to identify key areas for improvement that can be evaluated into more details with a different tool. |
| <u>Threats</u><br>This tool might be criticized because of | The tool is not well adapted to evaluation of complex surveillance systems that encompass multiple hazards and components, such as AMU and AMR, given that the surveillance of different AMR bacteria may differ in the same surveillance system.<br>If results of evaluation or its application are not discussed with key people, its simplicity may lead to a superficial evaluation of certain aspects.<br>Some indicators are not applicable to country or program context e.g., added value of OH integration in the case a system was integrated from its beginning.<br>Because data collection is expected to be short (e.g., no interviews), it is critical to have the right experts around the table to provide the required knowledge.<br>Not suitable for end-users of the system.<br>To ensure full comprehension of some indicators, previous clarification of their aim may be required, giving special attention to the terminology used, before conducting a meeting with relevant stakeholders.<br>While the tool provides output figures describing the level of OH-ness, it does not allow to visualize the actual system (distribution of surveillance programs by sector and domain) or collaboration between actors/programs (e.g., via social network analysis). Adding this feature would be an asset. |

### 3.1 Functional aspects

Regarding ease-of-use, the tool scored highly mainly due to its user-friendly interface with checkboxes to answer the indicators. When it came to the scope, the OH-EpiCap tool does not seem to cover all aspects connected with OH surveillance.

As for prerequisites to use the tool, no previous data collection is required, and the answers can be given based on the evaluators’ experience connected with the surveillance activity. Most indicators require that the evaluation is conducted with specialists in the surveillance activity, or that they are consulted in the process, given that an in-depth perspective of the whole surveillance system is needed. No training is necessary to get acquainted with the tool, however it is recommended that at least one of the evaluators gets acquainted with the indicators and clarifies any doubts before organizing a workshop with the specialists involved in the evaluation and other relevant actors.

The tool is free to use, and it can be successfully applied by a small group, involving three to four persons in most cases, providing that the group can form a clear cross-sectoral picture of the surveillance system. Depending on the expertise of the stakeholders gathered, the evaluation can be conducted in half a day or slightly longer.

The outputs generated provide an excellent overview of the responses given. However, the content of the evaluation needs to be discussed with relevant actors before it can be translated into specific changes in the surveillance activity.

### 3.2 Content themes

The tool does not encompass indicators specifically addressing AMU and AMR surveillance. Even though not covered to a complete extent, the OH-EpiCap provides an excellent overview of the thematic areas connected with the human and budget resources needed to maintain the surveillance activity, as well as the collaboration in the governance structures of the system and in the technical surveillance activities. It also encompasses indicators about the possible adaptation of the surveillance activities to new challenges and in an efficient manner. The overall impact of the surveillance activity is also covered, but the tool does not go into details regarding how the information generated by the surveillance activity could lead to changes in the health outputs. It also does not go into details in the governance domain, specifically the accountability of stakeholders, the coordination of activities and the transparency of processes.

## 4 Discussion

### 4.1 Overall perception on the tool

According to the information collected in the nine case studies, the OH-EpiCap can provide an overview of several crucial topics connected with AMR integrated surveillance, even though the tool is not specifically designed to evaluate these activities. The OH-EpiCap tool provides a superficial assessment of the three dimensions targeted, which cover most of the elements that are important for assessing surveillance systems, as described in the Integrated surveillance systems evaluation (ISSE) framework (5). The ISSE framework identified five levels of assessment for such surveillance systems, which include the integration of a OH approach, the production of integrated information and expertise, the generation of actionable knowledge, the influence on decision-making and the contribution to desirable outcomes. Evaluating these five levels in a comprehensive manner requires important time and resources, and the OH-EpiCap tool constitutes a good first step towards their overall evaluation.

Simplistic design and user friendliness, without requiring training of evaluators are highly appreciated, not just by our assessors but also among users in general as shown in a survey recently undertaken among surveillance program practitioners and evaluators (17).

The outputs generated by the OH-EpiCap may not lead directly to actions, however these can provide the basis for discussing further improvements with relevant stakeholders, as presented in a case study by Moura et al. (2022). The MATRIX project also encompasses other activities that are complementary to the OH-EpiCap, such as the “Roadmap to develop national One Health Surveillance” which aims to function as a guideline for the development of OH Surveillance activities according to needs and resources in different countries (18).

An evaluation using the OH-EpiCap can be conducted in a short period of time and with a small group of stakeholders, making it feasible to conduct an evaluation in situations with low resources and recurrently, when changes are implemented, benchmarking the system with itself over time. This can be made easily as the OH-EpiCap tool contains benchmarking functionalities. These functionalities were not investigated in the present study, because of the different aims and purposes of the systems evaluated as noted in Table 2, e.g., the Danish DANMAP serves the purpose of integrated monitoring of AMU and AMR for both the animal and human sectors, while the Italian ClassyFarm encompasses mainly farm-level risk categorization components (e.g. biosecurity and animal welfare, besides AMR and AMU) whereas the AMR surveillance in the human sector is conducted by different institutions (19). Both approaches are valuable for AMR surveillance and control and ultimately connected with the objectives of the country, but the approaches are not comparable. It is also worth mentioning that, given the above-mentioned differences in the aims of the surveillance activities evaluated, questions connected to real-time response capacity were considered not relevant in certain cases.

### 4.2 Recommendations

AMR surveillance systems are complex and encompass multiple hazards e.g., surveillance of clinical isolates in human health, bacterial isolates from animals at slaughter lines and in slurry and sewage systems, each with their own particularities and logistics (5). So, an overall picture had to be thought of to answer some indicators, which may justify the application of the tool to several of these components, focusing on one hazard at a time.

Most of the indicator questions were considered simple and straight forward. However, considering the expected worldwide application of the tool by users, who may have different use of the English language and, hence, familiarity with the terminology used, materials should be developed to unequivocally clarify the aim of all indicators. With the publication of case studies evaluations and the scientific paper accompanying the tool (14), this should be accounted for. It is also important to note at the time of writing this work, the OH-EpiCap tool was still in a Beta Version, so the phrasing of indicators was not final. The tool was subsequently applied in nine different countries, by different native language users, providing important feed-back to the developers regarding the phrasing of the indicator question.

### 4.3 Possible application of the OH-EpiCap

By highlighting components, which may be improved in a hazard integrated surveillance activity, the OH-EpiCap is a valuable new addition, which can act as a simple gateway to conduct a more in-depth evaluation of certain surveillance system components as considered relevant. This may be done by using other preestablished tools designed to evaluate OH integration, such as the Evaluation of Collaboration for Surveillance (ECoSur) or The Network for Evaluation of One Health (NEOH).

ECoSur has been developed to facilitate an in-depth analysis of the organization and functioning of collaboration taking place in a multisectoral surveillance system, aiming to evaluate the overall quality and relevance of such collaboration in meeting the objectives envisioned by stakeholders to produce the expected outputs of the program (20). From a user’s perspective, this tool gives a detailed evaluation of multisectoral collaboration in OH surveillance activities, however it requires a high level of abstraction to understand the indicator questions listed in the tool. Moreover, conducting a full evaluation is rewarding regarding quality of output, but remains time and resource demanding (12).

NEOH allows the evaluation of the coherence between operational and organizational aspects of OH activities, with the aim of identifying the added value of the integration across disciplines and sectors (21). From a user’s perspective, this tool is a comprehensive, multi-faceted fit for a transversal and detailed analysis of OH initiatives. However, conducting an evaluation using NEOH may be difficult and time consuming given that users should have specific training in systems thinking to make the most out of it (12).

One of the ongoing activities in the CoEvalAMR network aims to simplify the application of the NEOH and ECoSur tools, using a modular approach. Given the complexity of evaluating integrated antimicrobial resistance surveillance systems, this could be of great value, targeting the evaluation to certain components which need to be prioritized.

Within the CoEvalAMR network, case studies have already been conducted from a user’s perspective on the application of the ECoSur (22) and the NEOH (23–26) tools. Other tools and frameworks that have been specifically designed to evaluate integrated AMR surveillance have also been evaluated: the FAO Progressive Management Pathway for AMR (FAO-PMP-AMR) (27–30) designed to guide countries in the implementation of National Action Plans for AMU and AMR (31); the FAO Assessment Tool for Laboratories and AMR Surveillance Systems (FAO-ATLASS) (32) developed to facilitate the assessment and definition of targets to improve national AMR surveillance systems in the food and agriculture sectors (33) and the Integrated Surveillance System Evaluation framework (ISSE framework) (34,35) developed to structure an assessment of the added value of integration in AMR surveillance systems (36). The selection tool developed by the CoEvalAMR network can help users to select an appropriate tool for their needs (37).

## 5 Conclusion

The OH-EpiCap is a welcome new addition to the portfolio of existing tools to evaluate integrated AMR surveillance systems. It provides a brief macro-overview of relevant OH topics, such as the perceived added value of establishing a OH team as a governance structure, serving as a basis to discuss possible adaptations of AMR surveillance activities, or targeting areas that may be further investigated using other established tools. It is easy to use, requires no training, and can be performed in less than a day, on the condition that detailed knowledge about the surveillance system to evaluate is present in the group performing the evaluation.

## Data Availability

The original contributions presented in the study are included in the article/supplementary material,
further inquiries can be directed to the corresponding author/s.

## 7 Author Contributions

The main author drafted the first version of the paper together with LA and LC, which was then commented by all authors. All authors read and approved the final version of the manuscript.

## 9 Acknowledgements

The authors acknowledge Henok Ayalew Tegegne, Carlijn Bogaardt, Lucie Collineau, Géraldine Cazeau, Renaud Lailler, Johana Reinhardt, Emma Taylor, Joaquin M Prada, Viviane Hénaux for the development of the OH-EpiCap tool and for making it available to conduct this work, as well as the stakeholders who provided information to conduct the evaluations in the different case studies.

