## Supplementary table 1 & supplementary figure 1 for "Users’ perception of the OH-EpiCap evaluation tool based on its application to nine national antimicrobial resistance surveillance systems"

### 1. Supplementary material

Table S1: Description of way of working when assessing the OH-EpiCap tool

| Country | Number of assessors and affiliation | Type of workshop | Total duration of the evaluation |
| --- | --- | --- | --- |
| Bangladesh | Academia (n=1) | One virtual meeting | 3 hours |
| Belgium | Academia (n=1) and public health agency (n=2) | One face-to-face meeting | 4 hours |
| Canada | Academia (n=2) | One virtual meeting | 8 hours -   The evaluation was based on the assessors' knowledge and previous data collection: two sessions of group discussions with the surveillance system stakeholders (duration = 4 hours) |
| Denmark | Surveillance system management (n=2), academia (n=2), and livestock industry (n=1) | Two virtual meetings | 4 hours |
| France | Public health agency (n=1), food safety agency n (=1), Ministry of Agriculture (n=1) | One virtual meeting* | 6 hours -   Data collection was facilitated by a previous case based on the application of ECoSur to the French AMR surveillance system and involving the realization of 52 semi-directed interviews. |
| Italy | Academia (n=2) | Two virtual meetings | 6 hours -   The evaluation was also based on assessors' knowledge, gained in previous meetings with the ClassyFarm stakeholders to evaluate the surveillance system, using NEOH and FAO-PMP tools. |
| Norway | Academia (n=3) | One round virtual meeting*  * After one round with only one assessor for familiarisation of the tool | 2 hours - The information used was based on the assessor's knowledge as they have long experience, being working on the surveillance of AMR in Norway for more than 20 years. |
| Portugal | Surveillance system management/academia (n=2) | Online discussion | 4 hours |
| United Kingdom | Academia (n=1) | Online discussion | 3 hours -   The information used was based on the assessor’s knowledge from previous work and data collection conducted, so no further data collection was needed. |

**Figure S1**: Example of a visual representation generated by the OH-EpiCap tool

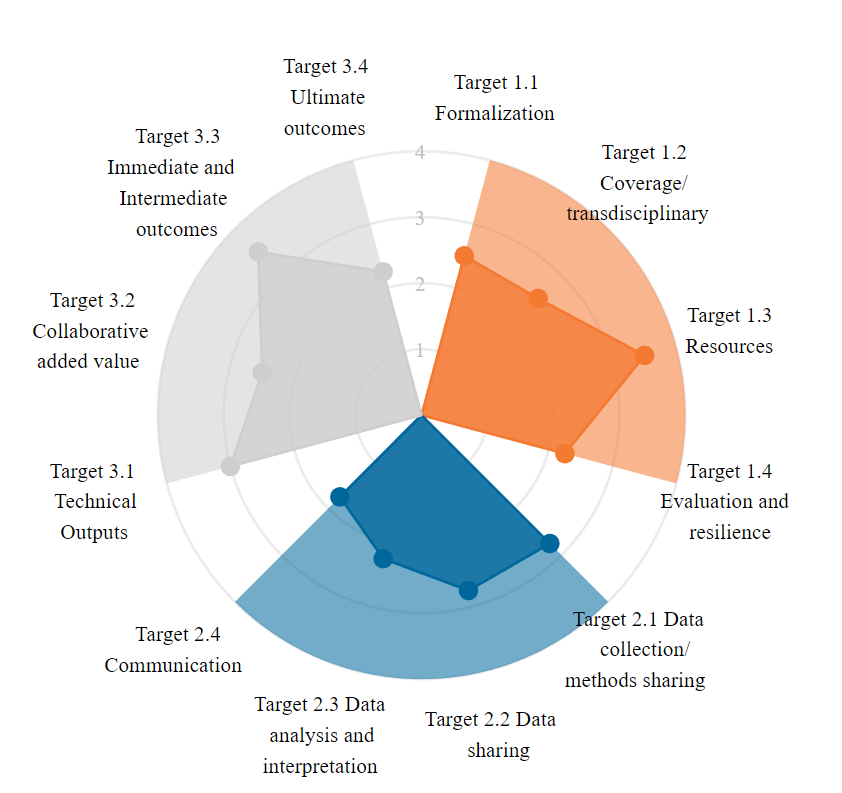
